# Identification and Analysis of Shared Risk Factors in Sepsis and High Mortality Risk COVID-19 Patients

**DOI:** 10.1101/2020.05.05.20091918

**Authors:** Sayoni Das, Krystyna Taylor, Matthew Pearson, James Kozubek, Marcin Pawlowski, Claus Erik Jensen, Zbigniew Skowron, Gert Lykke Møller, Mark Strivens, Steve Gardner

**Affiliations:** PrecisionLife Ltd, Long Hanborough, Oxford, UK

## Abstract

**BACKGROUND:** Coronavirus disease 2019 (COVID-19) is a novel coronavirus strain disease caused by severe acute respiratory syndrome coronavirus 2 (SARS-CoV-2). The disease is highly transmissible and severe disease including viral sepsis has been reported in up to 16% of hospitalized cases. The admission characteristics associated with increased odds of hospital mortality among confirmed cases of COVID-19 include severe hypoxia, low platelet count, elevated bilirubin, hypoalbuminemia and reduced glomerular filtration rate. These symptoms correlate highly with severe sepsis cases. The diseases also share similar comorbidity risks including dementia, type 2 diabetes mellitus, coronary heart disease, hypertension and chronic renal failure. Sepsis has been observed in up to 59% of hospitalized COVID-19 patients.

It is highly desirable to identify risk factors and novel therapy/drug repurposing avenues for late-stage severe COVID-19 patients. This would enable better protection of at-risk populations and clinical stratification of COVID-19 patients according to their risk for developing life threatening disease.

**METHODS:** As there is currently insufficient data available for confirmed COVID-19 patients correlating their genomic profile, disease severity and outcome, co-morbidities and treatments as well as epidemiological risk factors (such as ethnicity, blood group, smoking, BMI etc.), a direct study of the impact of host genomics on disease severity and outcomes is not yet possible. We therefore ran a study on the UK Biobank sepsis cohort as a surrogate to identify sepsis associated signatures and genes, and correlated these with COVID-19 patients.

Sepsis is itself a life-threatening inflammatory health condition with a mortality rate of approximately 20%. Like the initial studies for COVID-19 patients, standard genome wide association studies (GWAS) have previously failed to identify more than a handful of genetic variants that predispose individuals to developing sepsis.

**RESULTS:** We used a combinatorial association approach to analyze a sepsis population derived from UK Biobank. We identified 70 sepsis risk-associated genes, which provide insights into the disease mechanisms underlying sepsis pathogenesis. Many of these targets can be grouped by common mechanisms of action such as endothelial cell dysfunction, PI3K/mTOR pathway signaling, immune response regulation, aberrant GABA and neurogenic signaling.

**CONCLUSION:** This study has identified 70 sepsis related genes, many of them for the first time, that can reasonably be considered to be potentially relevant to severe COVID-19 patients. We have further identified 59 drug repurposing candidates for 13 of these targets that can be used for the development of novel therapeutic strategies to increase the survival rate of patients who develop sepsis and potentially severe COVID-19.

## Introduction

The 3.5 million confirmed COVID-19 cases worldwide have resulted in 250,000 deaths up to 4 May 2020^1^. While around 80% of cases will result in mild forms of the disease, a study of admissions in 166 UK hospitals showed that around 33% of all patients requiring hospitalization and 45% of those requiring high dependency or intensive care have died from the disease^2^. There is a significant correlation with age, sex and underlying co-morbidities, with elderly males at highest mortality risk.

There has been some success in repurposing existing antiviral therapies such as the investigational agent Remdesivir in COVID-19 patients^3^, but antiviral agents tend to be more effective when applied early in the disease course. It would be highly desirable to understand the differential in host response between patients in terms of their underlying genetic, epidemiological and clinical factors to be able to predict which individuals are more at risk of developing severe COVID-19, and to identify potential therapies for the later stages of the disease, which are driven by the host’s immune response rather than the coronavirus^4^.

Severe disease has been reported in up to 16% of hospitalized cases and often requires forced oxygen therapy and/or sedation and ventilation of patients. A viral sepsis has been observed in many patients and is thought to be crucial to the disease mechanism of COVID-19^5^. The admission characteristics associated with increased odds of hospital mortality among SARS-CoV-2 confirmed cases of COVID-19 include severe hypoxia, low platelet count, elevated bilirubin, hypoalbuminemia and reduced glomerular filtration rate^6,7^. These are symptoms that are also highly correlated with severe sepsis cases. The two diseases also share similar co-morbidity risks including dementia, type 2 diabetes mellitus, coronary heart disease, hypertension and chronic renal failure^8^. Sepsis has been observed in up to 59% of hospitalized COVID-19 patients^9^.

Sepsis is itself a life-threatening condition with a mortality rate of 20% that can result from a patient’s dysregulated systemic inflammatory response to a pathogen^10^. It is one of the leading causes of mortality in hospitals in the US, representing a major socioeconomic burden^11^.

In sepsis, there is an intersection between the inflammatory and hemostatic pathways, with the simultaneous activation of both the inflammatory and the coagulation cascades. The spectrum of this interaction can vary from mild thrombocytopenia to fulminant disseminated intravascular coagulation (DIC)^12^. Perhaps due to the heterogeneous nature of the disease, very few GWAS have found significant genetic loci relating to sepsis risk^13^. Of these genes, the majority relate to endothelial cell and immune response pathways but there is still very little known about the genetic risk factors underlying susceptibility to developing sepsis.

While sepsis in itself represents a significant area of unmet medical need, the studies cited above provide evidence that many of the patients who developed a severe response to SARS-CoV-2 also presented with sepsis. Having identified a significant number of risk-associated genes in a sepsis population, we aim to investigate if those risk variants are also present in patients who have been hospitalized with severe COVID-19. If so, this would provide insights into the shared risk factors and potential novel therapeutic options for both diseases.

## Methods

We analyzed a dataset using sepsis patients found in the UK Biobank^14^, containing sepsis patients (n=6,843, 3,700 males and 3,143 females) and age and co-morbidity matched controls (n=6,820, 4,295 males and 2,525 females). Due to limited data availability, it was not possible to fully sex match the controls. The available genotype data for the cohorts included 542,245 SNPs after quality control. Cases were identified using the ICD-10 codes relating to hospital admission with sepsis following infection from a number of different pathogenic sources. The case criteria included a series of seven septicemia related ICD-10 codes (see Appendix for full details).

The construction of an appropriate control set is more challenging with an infectious disease as the lack of disease may result from lack of exposure as much as a genetic protective effect or other predisposition to resistance. To overcome this, we included only patients who had been exposed to some of the most common sepsis-causing pathogens (such as *Staphylococcus* and *Streptococcus* species) but not gone on to develop sepsis. The control group criteria also included at least one of the ICD-10 codes for the most common chronic co-morbidities known to increase a patient’s risk of developing sepsis (including patients with diabetes, hypertension, chronic renal and/or liver disease). We then ranked the controls who met these criteria by age, selecting the oldest first, as age is also another critical phenotypic risk factor for sepsis. Controls were therefore selected for maximum risk and exposure, and age matched.

One limitation of the UK Biobank dataset is that the ethnicity distribution of the participants is heavily skewed to white British participants and it has consequently not been possible to fully investigate additional risk factors in BAME patients.

The case and control population co-morbidity profile is shown in Figure 1.

**Figure 1:**
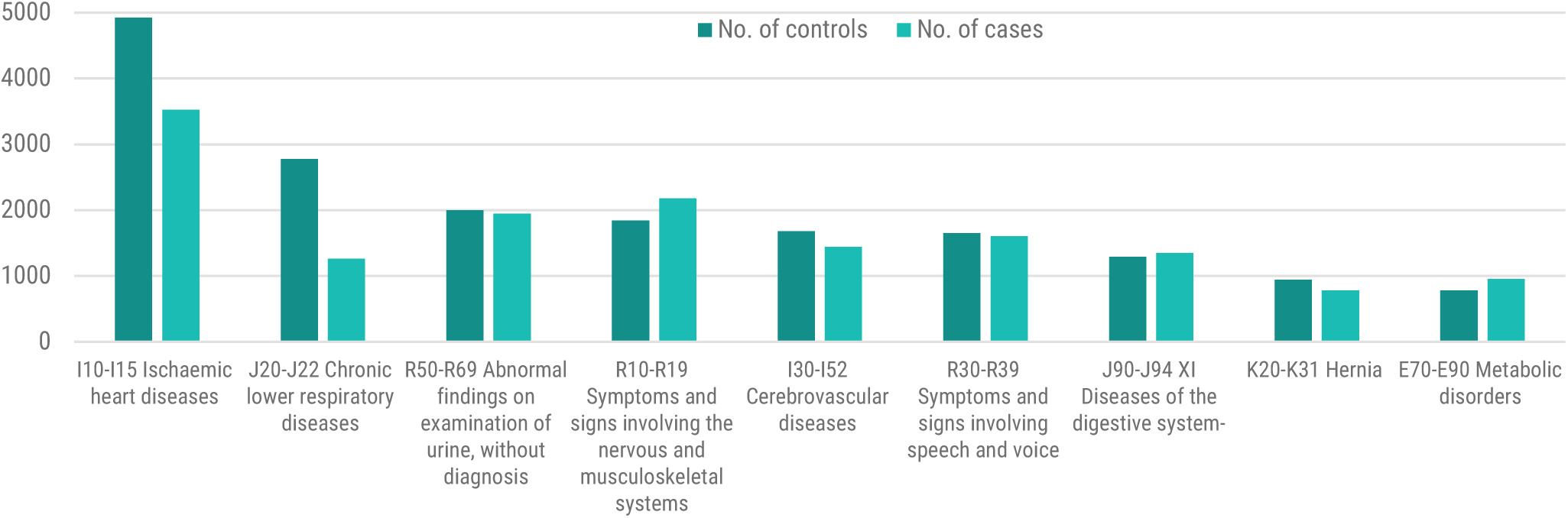
Incidence of co-morbidities by ICD-10 code from UK Biobank in the sepsis dataset (n=13,663 split 6,843 cases and 6,820 controls)

**Figure 2:**
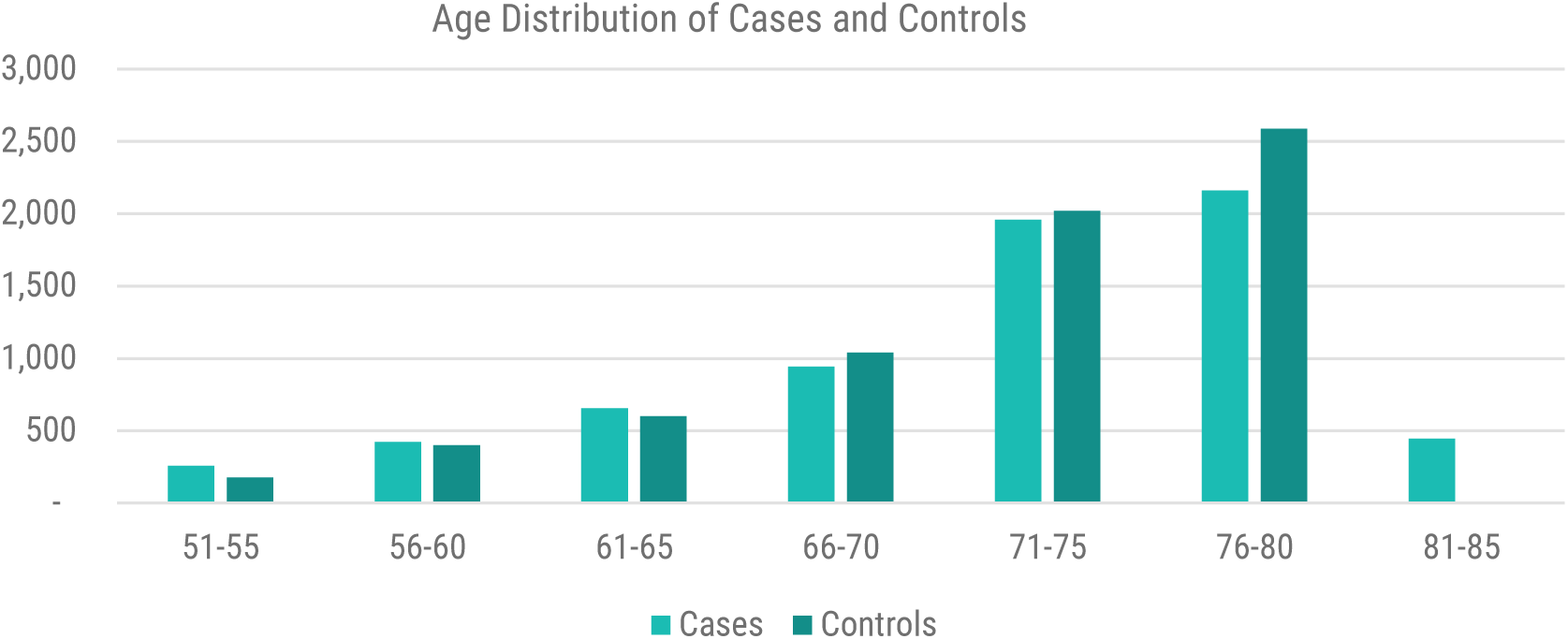
Age distribution of cases vs controls from UK Biobank in the sepsis dataset

We used the **precision**life combinatorial multi-omics platform to identify sepsis associated SNPs and genes from the sepsis case:control dataset. **precision**life is a multi-omics (genomic, proteomic, transcriptomic, phenotypic) association platform that enables the hypothesis-free detection of high order disease associated combinations of features (e.g. SNP genotypes) at genome wide study scale^15^. It finds and statistically validates combinations of features (typically three to ten features in combination known as ‘signatures’) that together are strongly associated with a specific disease diagnosis or other clinical phenotype (e.g. fast disease progression or therapy response). It can combine genomic, transcriptomic and epidemiological data in its analyses. When used to analyze genomic data from patients, **precision**life can identify high-order epistatic interactions comprising multiple consistently co-associated SNP genotypes. This analytical mining platform has been validated in multiple disease populations^16,17^.

The analysis and annotation of the sepsis associated combinatorial genomic signatures in this dataset using the **precision**life platform took less than a day, working hypothesis-free, running on a dual CPU, 4 GPU compute server.

The sepsis disease signatures identified by the analysis were then annotated and disease-associated SNPs were mapped to the human reference genome^18^ in order to identify disease-associated and clinically relevant target genes. A semantic knowledge graph derived from multiple public and private data sources was used to annotate the SNPs and targets, providing sufficient contextual information to test the targets against the 5Rs criteria of early drug discovery^19^ and forming strong, testable hypotheses for their mechanism of action and impact on the sepsis disease phenotype.

We also applied a series of heuristics to the resulting sepsis risk-associated genes in order to identify novel sepsis targets with high potential either for drug development or drug repurposing. These criteria include a strong association of the target with the disease and its pathophysiological mechanisms, relevant tissue expression, tractability for novel disease targets amongst several others. Additional criteria used for prioritizing repurposing targets included known drugs, patent scope, favorable pharmacokinetic and toxicology profiles for repurposing targets.

We then sought to correlate these findings with COVID-19. From the UK Biobank, we currently have genotype data for 572 patients who have tested positive for COVID-19 in hospital and 97 patients who have tested positive but have not been hospitalized. Although there is as yet no additional clinical data available on these patients, given the way in which UK patients have been admitted and treated, we believe that hospitalization for the disease could be used as a surrogate indicator of having a relatively severe form of the disease. We compared the significant disease associated combinatorial signatures found in the sepsis cohort against these COVID-19 patients, in order to investigate if any of them are also present in the COVID-19 population.

## Results

Conducting a standard PLINK^20^ GWAS analysis on the UK Biobank sepsis dataset that we generated revealed no significant SNPs (Figure 3). This is not an unexpected result as it replicates the findings of existing sepsis GWAS studies, which have yielded few results previously^21,22^. It is also consistent with the initial findings reported by the COVID-19 Host Genetics Initiative^23^ based on data from 917 COVID-19 cases.

**Figure 3:**
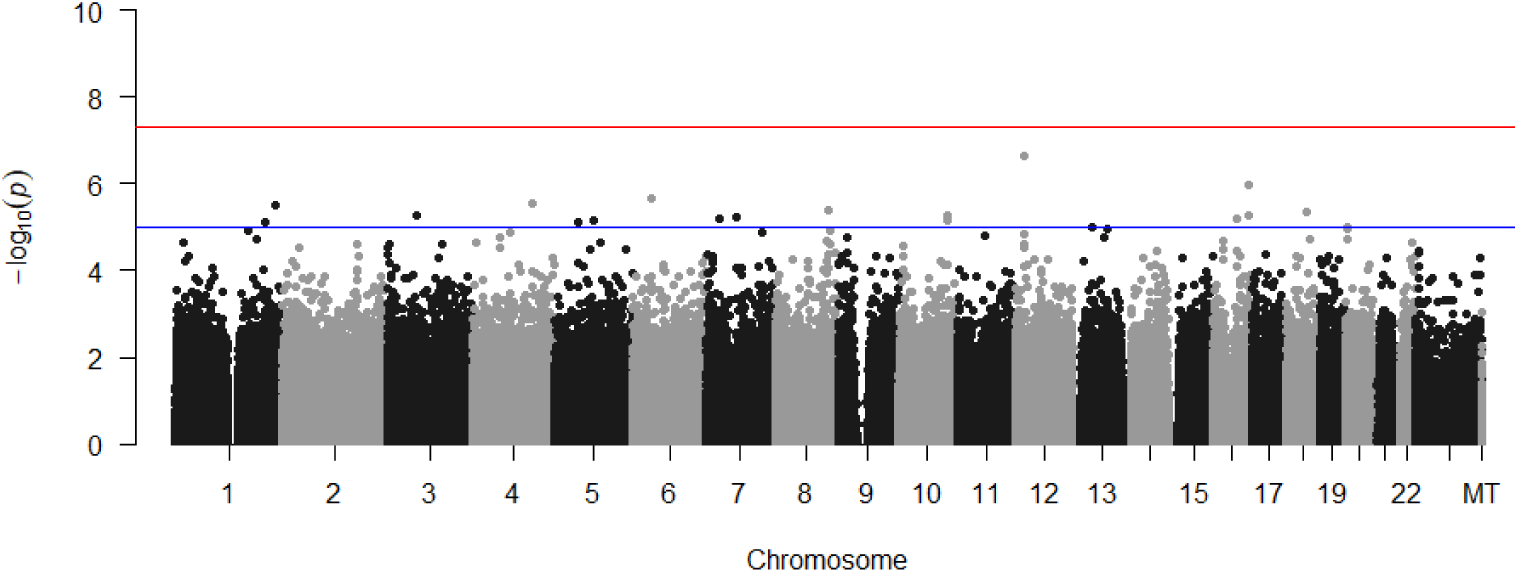
Manhattan plot generated using PLINK of genome-wide p-values of association for the UK Biobank sepsis dataset. The horizontal red and blue lines represent the genome-wide significance threshold at p<5e-08 and p<1 e-05 respectively.

In contrast with the single SNP association basis of PLINK, the **precision**life platform constructs high-order networks of co-associated SNPs (signatures), which are associated in combination with the disease phenotype. Some of the component SNPs in these signatures will fall below the GWAS p-value threshold when evaluated individually across the whole population. However, in combination, the signatures can be proven to be highly significant using multiple discriminative statistical techniques.

Running the same sepsis dataset using the **precision**life platform, we identified 1,446 combinations of SNP genotypes (disease signatures) representing different combinations of SNP genotypes within the sepsis patient population (Table 1). These signatures consisted mainly of 3 to 5 SNP genotypes in combination, where most of their constituent SNPs would not have been discovered using standard GWAS analyses.

**Table 1:** Summary of **precision**life sepsis disease study run and results. Number of validated N-states, simple networks and critical SNPs identified in the sepsis study using a 5% False Discovery Rate and 250 cycles of fully random permutation.

|  | **Sepsis Study** |
| --- | --- |
| **Dataset** (542,245 SNPs) | 6,843 cases<br>6,820 controls |
| **False Discovery Rate (FDR)** | 5% |
| **N-states** | 1,446 |
| **Simple networks** | 145 |
| **No. of unique SNPs in merged network** | 1,904 |
| **RF scored SNPs** | 136 |
| **Prioritized Genes** | 70 |
| **Penetrance**<br>(cases containing critical SNPs from the disease N-states) | 3,781 (55%) |

All of the SNP genotypes and their combinations were scored using a Random Forest (RF) algorithm based on a 5-fold cross-validation method to evaluate the accuracy with which the SNP genotype combinations predict the observed case: control split. The majority of the critical SNPs were assigned a high RF score by the algorithm, thus confirming that they have a strong association with the disease (Figure 4A). The chromosomal distribution of the critical SNPs is shown in aggregate in Figure 4B and in detail in Figure 4C.

**Figure 4:**
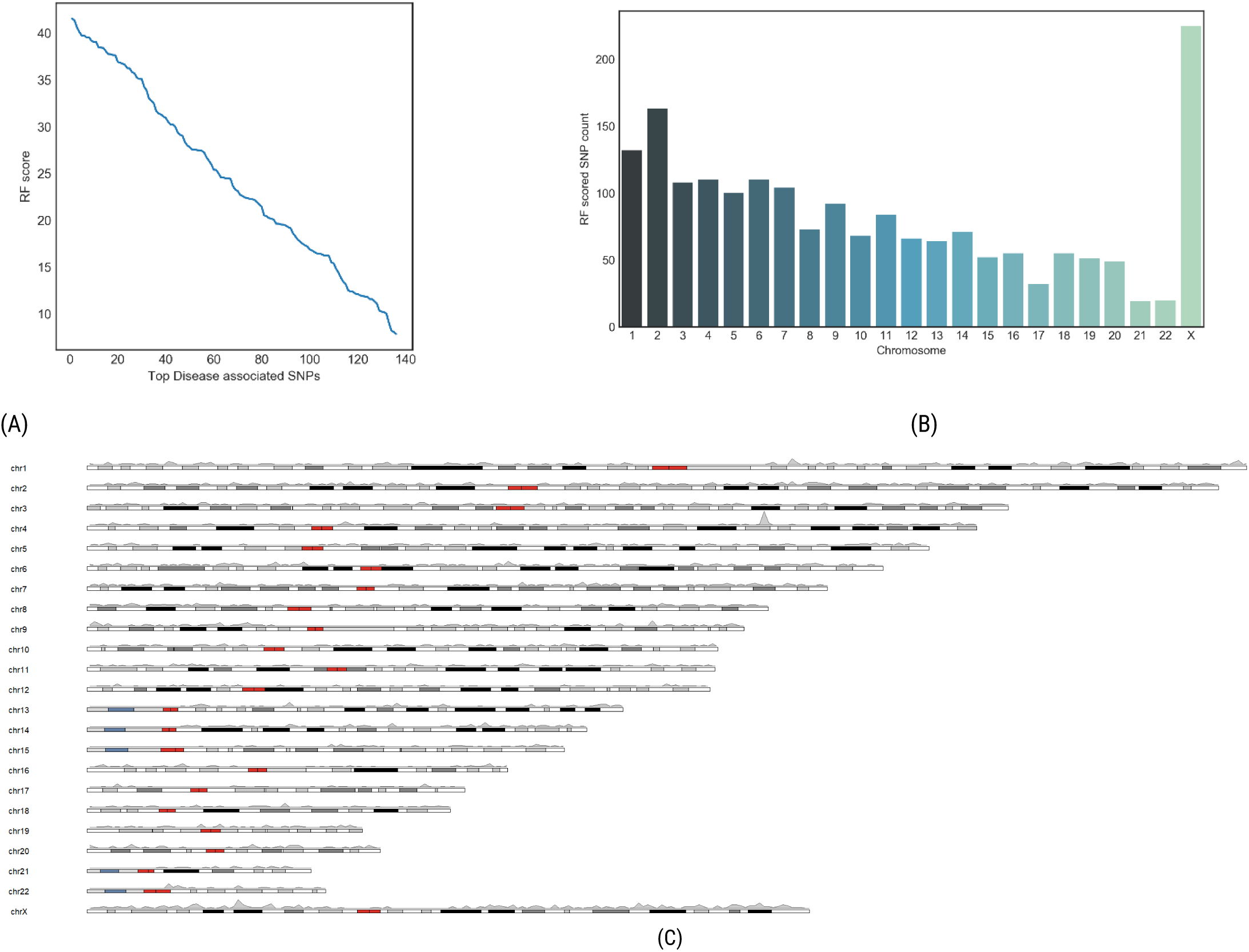
Distributions of (A) RF scores and (B) and (C) chromosomal locations for critical disease associated SNPs

These SNP networks were clustered based on their co-occurrence in patients, which generated a complex disease architecture view showing the distinct patient sub-populations (Figure 5). The most critical (and predictive) SNP genotypes found at the center of the SNP networks shown in Figure 5 are found in 3,781 of the 6,843 cases (55%).

**Figure 5:**
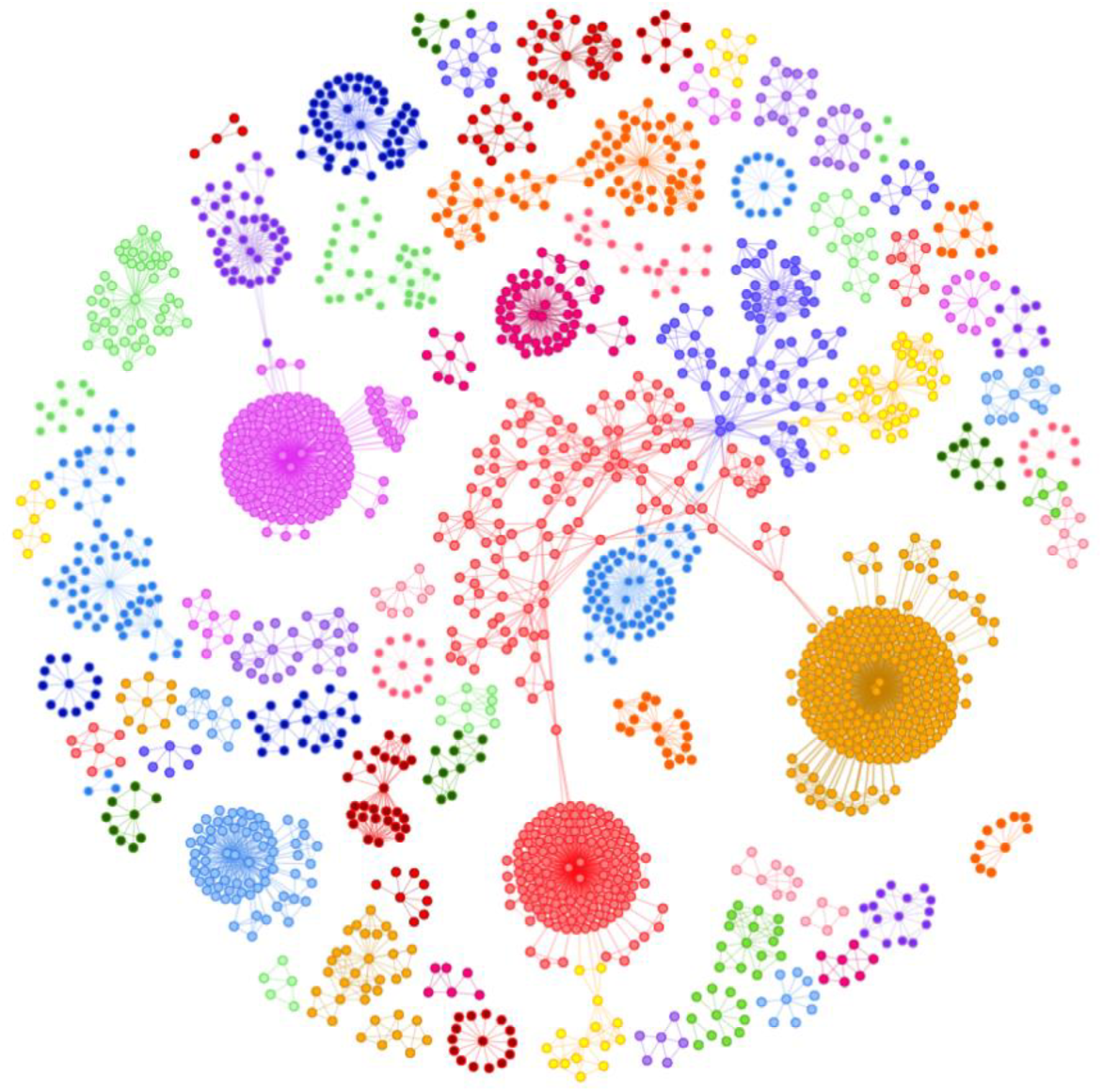
Disease architecture of the sepsis cohort generated by the **precision**life platform. Each circle represents a disease associated SNP genotype, edges represent co-association in patients, and colors represent distinct patient sub-populations or ‘communities’.

Mapping the highest-scoring SNPs to genes revealed 70 genes that are strongly associated with the risk of developing sepsis. Of these genes, several have already been shown to be implicated in sepsis pathogenesis in the scientific literature, providing validation for the hypothesis-free combinatorial approach to analyzing complex disease populations. In contrast, a recent meta-analysis reviewing all previous studies investigating genetic variants and sepsis risk found just 23 genes that were significantly associated with disease susceptibility^24^.

Functional enrichment analysis of the highest-scoring genes indicated that a significant number of them are involved in processes such as stress response, leukocyte and immune system activation, cell adhesion and autophagy (Figure 6).

**Figure 6:**
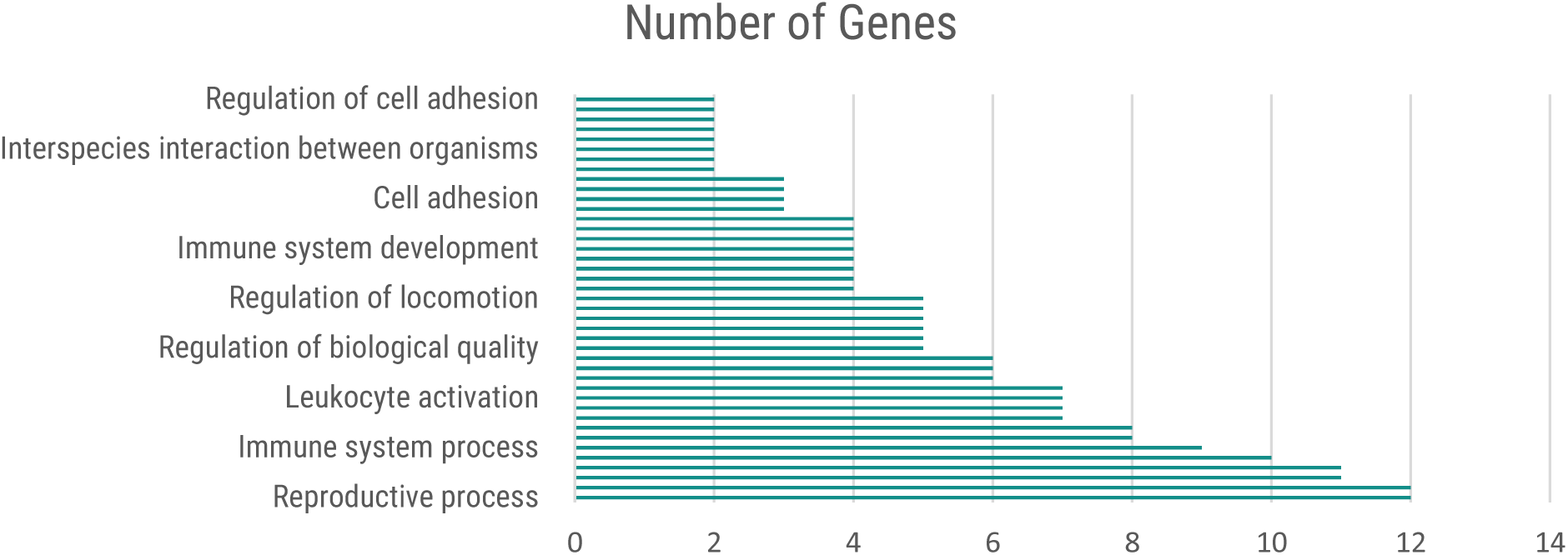
Functional categories of genes defined by high-level Gene Ontology terms using ShinyGo.

Of the 70 sepsis risk genes we identified, 13 of them are targeted by active chemical compounds. Seven of those genes are targeted by licensed drugs and could therefore represent potential drug repurposing opportunities. The specific genes and pathways are described in the Appendix and some are outlined further in the Discussion section.

As shown in Figure 4B, there is a significant cluster of SNPs and genes associated with the X chromosome. It may be a coincidence that some of the key genes for COVID-19 infection (e.g. *ACE2* and angiotensin II) are located on this chromosome, and that by having two copies of the X chromosome women might be offered some level of protection against the effects of the disease. There is a small possibility that it may also be related to the lack of complete sex matching of the controls. This observation will be the subject of further study.

**Figure 7:**
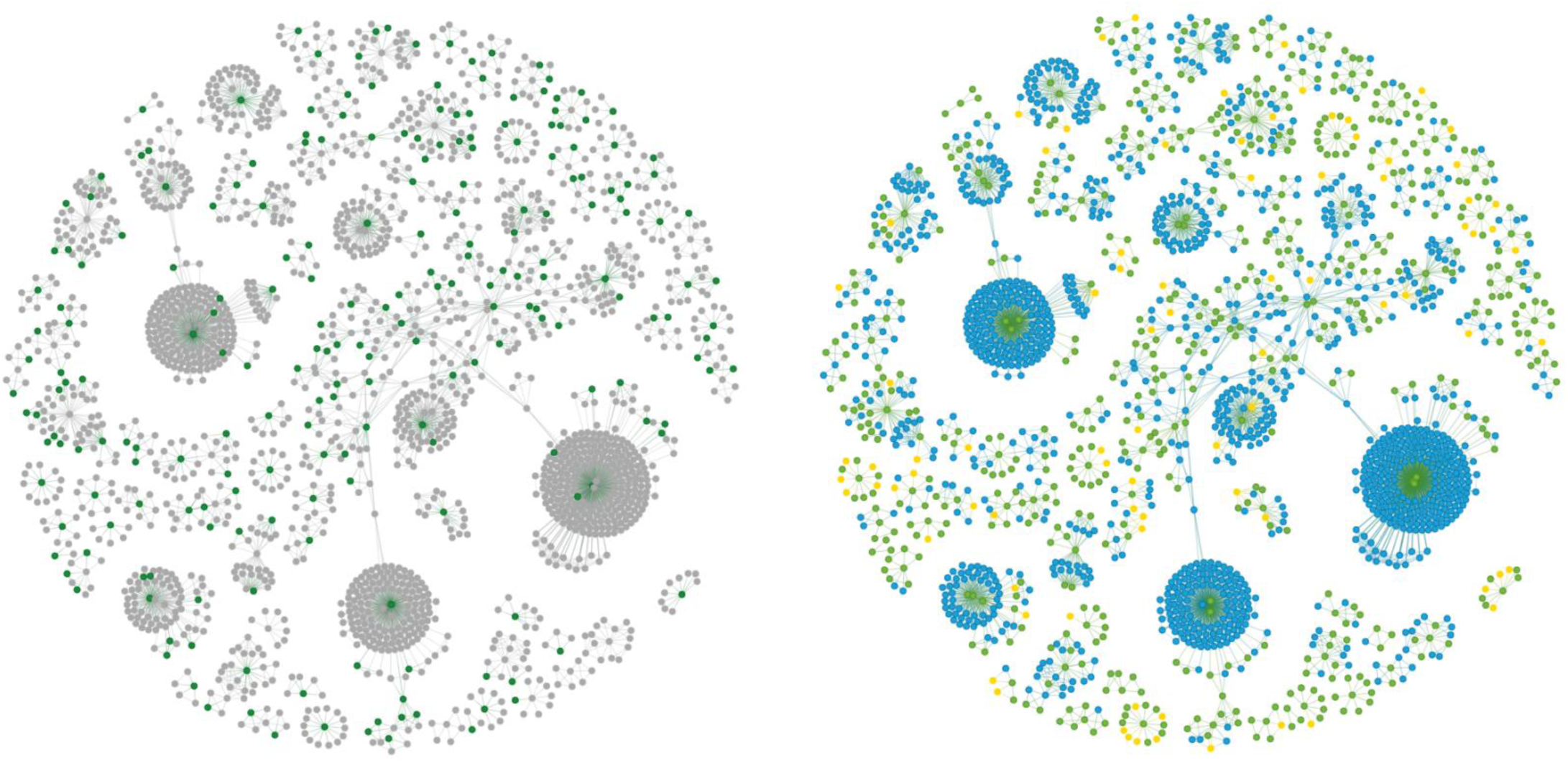
Disease architecture of the sepsis cohort highlighting the SNPs found on the X chromosome (left in green) and showing SNP genotypes (right - homozygous major allele = blue, heterozygous = green, homozygous minor allele = gold).

We focused our current analysis on the autosomal SNPs and searched for sepsis associated combinatorial signatures in 572 patients who were hospitalized and tested positive for COVID-19. 51 of the 572 patients have at least one of the significant N-states found in the sepsis population, indicating an element of autosomal genetic overlap between patients who are at risking developing sepsis and those who develop severe reactions to SARS-CoV-2. In total, 33 sepsis disease signatures were found in these patients that were not observed in COVID positive patients showing mild symptoms or any sepsis controls. We have described the likely functional impact of one of these disease signatures in the Discussion section.

## Discussion

Many of the risk-associated genes found using the **precision**life platform can be grouped into one of four common biological pathways that can be linked to sepsis pathogenesis; immune-related genes, CNS signaling molecules, the PI3K/mTOR pathway, and targets relating to the endothelium.

### PI3K/AKT/mTOR Pathway

Out of the 70 significant genes found, 21 of them have already been investigated in the context of cancer, ranging from *in vitro* and preclinical studies through to large clinical trials. The most common pathway these targets centered around was the PI3K/Akt/mTOR pathway, which is implicated in driving oncogenesis through its effects on cell proliferation and apoptosis^25^.

We identified genes that are direct regulators of PI3K (phosphoinositide-3-kinase). Mutations in these classes of genes are associated with immune deficiency syndromes, resulting in chronic viremia, recurrent respiratory tract infections and defects in B cell development^26,27^. PI3K plays a key role in both the adaptive and innate immune response, as it plays a critical role in the development of regulatory natural killer (NK), memory T and B lymphocytes, as well as functional phagosome formation^28^. It is also crucial for the activation of the IL-12 negative feedback pathway that helps to resolve inflammation. In this way, we can hypothesize that the PI3K pathway is critical for regulating the immune response during the development of sepsis. This theory is supported by a study that demonstrated that activation of PI3K/Akt signaling attenuated pro-inflammatory markers, neutrophil infiltration and apoptosis in a model of sepsis-induced cardiac dysfunction^29^.

However, there is some contradictory evidence surrounding the role of the PI3K pathway in sepsis, exemplified by a model of sepsis-induced myocardial dysfunction that showed selective inhibition of a PI3K isoform reduced the production of pro-inflammatory cytokines and other pathological markers of sepsis^30^. This indicates that PI3K also plays a crucial role in the regulation of the immune system in the context of sepsis pathogenesis.

We found two genes associated with mTOR (mammalian target of rapamycin) signaling. The mTOR pathway is a critical regulator of T-cell differentiation and function, with inhibition of mTOR activity resulting in increased autophagy, and sustained development of memory CD8+ cells and regulatory CD4+ cells^31^. Furthermore, a study found that patients with sepsis had higher serum levels of key molecules in the mTOR signaling pathway^32^, indicating that high mTOR activity may play a role in driving key disease processes involved in sepsis.

In addition to this, we also identified three genes that indirectly affect the PI3K/Akt/mTOR pathway through interaction with PTEN, one the main inhibitors of this pathway^33^.

Our results indicate that whilst these apoptosis-related pathways are most commonly associated with oncogenesis, these cellular processes are also highly important in the regulation of the immune system and variants in these genes may also predispose patients to inflammatory diseases such as sepsis as well as cancer.

### Immune Response Genes

As to be expected, our analysis found significant genes relating to both the innate and adaptive immune response. The identification of over 10 genetic variants in a range of different immunological targets indicates that many patients who develop sepsis may have inherent aberrations in their immune response system when challenged by an infectious agent.

Of these targets, one gene encodes a T cell differentiation antigen that binds to key components of Gram-positive and Gram-negative bacteria, causing agglutination and inhibiting their virulence factors. Infusion of this CD receptor in a mouse model of sepsis resulted in improved survival and reduced systemic inflammation, indicating that patients with variants in this gene likely have defective pathogen binding abilities and greater susceptibility to infection by sepsis-causing bacteria.

We also identified several transcription factors involved in regulation of expression of certain classes of lymphocytes and MHC class II genes. One of these genes encodes a transcription factor that plays a role in regulating interferon-gamma induced genes, thereby modulating the duration of the inflammatory response, whilst a different gene we found is critical for class switch recombination (CSR) in activated B cells. Furthermore, we found a target encoding a transcription factor that is essential for the development of IL-17 producing T cells. High IL-17 is associated with a pro-inflammatory phenotype and increased risk of death in a mouse model administered with toxic shock syndrome toxin (TSS-1)^34^.

In addition to this, we identified a gene encoding a serine protease that inhibits the function of furin, an enzyme that cleaves a variety of different pathogenic substrates, including SARS-CoV-2 coronavirus^35^. Furin is upregulated during the adaptive immune response, enhancing the virulence of pathogens through the cleavage of key sites and allowing viral entry into the host cells^36^. Administration of a furin inhibitor in a mouse model of atherosclerosis attenuated vascular endothelial injury and pro-inflammatory cytokine expression, indicating that this could be a key target in several different facets of sepsis pathophysiology^37^.

### Endothelial Cell and Vascular Inflammation

Endothelial dysfunction is a well-established mechanism that contributes to sepsis pathogenesis^38^. A recent GWAS identified two genes (*FER* and *MAN2A1*) that are implicated in the endothelial response to pathogens. Our study identified four more targets that are associated with the vascular endothelium.

Two of these genes encode cell adhesion and ECM-interacting molecules that have been linked to endothelial inflammation and oxidative stress in models of atherosclerosis. One of these genes is inhibited by oxidized LDLs via the LOX-1 receptor, resulting in increased vascular inflammation and lipid accumulation. There is evidence that oxidized LDLs are upregulated in inflammatory conditions and inhibition of LOX-1 may help to attenuate the pro-inflammatory cascades seen in sepsis^39^.

A different risk-associated gene is regulated by angiotensin II and has been implicated in the promotion of smooth muscle fibrosis, oxidative stress and endothelial dysfunction in this way.

Furthermore, we have found a transcription factor that targets VEGF, a key regulator of angiogenesis and endothelial cell function. High levels of VEGF are observed in animal models of sepsis, where it potentiates the effects of TNF-α on endothelial cells and increases vascular permeability which is a key pathogenic event in severe sepsis^40^.

### Neuronal Signaling Mechanisms

Finally, we identified several variants in genes associated with neurogenic signaling pathways. The CNS plays a key role in regulating the immune system and cytokine expression, as well as the systemic vascular response observed in sepsis patients^41^.

One of these variants encoded a key enzyme involved in the production of GABA, the main inhibitory neurotransmitter in the CNS. Polymorphisms in this gene are associated with multiple neuropsychiatric disorders, including schizophrenia, epilepsy, bipolar, depression and anxiety. To the best of our knowledge, there is no evidence currently linking GABAergic signaling to sepsis, however studies have found increased levels of one of the targets that we identified in models of hippocampal inflammation and autoimmune conditions. Another target identified in our study encodes a G-protein-coupled receptor that regulates GABA_B_ neurons, indicating that polymorphisms affecting the function of genes involved in GABA signaling increase the risk of developing sepsis.

Alongside the GABA related targets this study identified, we also found targets in a number of other common neurotransmission pathways, including regulation (activation) of kainate-type glutamate receptors and the adenosine receptor, as well as four other genes that are key to neuronal development and synaptic plasticity.

One of the variants we found that was implicated in neurogenic signaling is located in *ADORA1*, which encodes for the adenosine A1 receptor. High levels of adenosine have already been found in the plasma of patients with septic shock and severe sepsis, indicating that it may play a key role in the development of the disease^42^. Furthermore, increased activation of adenosine A1 receptors in the lung results in increased Gram-negative-induced tissue injury through increased pro-inflammatory mediators^43^. A study has now shown that using adenosine A1 antagonists attenuates sepsis pathophysiology and improves survival in a rat model of the disease^44^.

### Drug Repurposing Opportunities

It is well-known that different diseases may share common pathways, and drugs that affect genes in these pathways may therefore be able to treat a variety of disease indications. The uniquely detailed disease architecture views generated by the combinatorial approach used in this study allow for the systematic repurposing of all known drugs against all the disease relevant targets identified. Mapping existing drugs onto the genetic and metabolic signatures identified for patient sub-groups indicates areas where there are already good clinical options, and also where trial use of existing therapeutics with good safety and tolerability profiles, with acceptable routes of administration, could have potential. For a given patient, their specific combination of SNPs will in large part determine which drug or combination of drugs are likely to benefit them personally.

We used the **precision**life platform to identify drug repurposing candidates for key disease-associated targets by mapping all of the existing drug options onto the genes found in different subgroups of the patient population. Applying a series of heuristics that we have developed we can rapidly identify candidates that may be therapeutically beneficial for select subsets of the patient population and to identify and efficiently prioritize those with the greatest repurposing potential for further investigation.

Of the 70 sepsis risk genes we identified, 13 of them are targeted by active chemical compounds found in DrugBank^45^ or ChEMBL^46^ and could therefore represent potential drug repurposing opportunities. In total there are 59 known active drugs identified for the 13 targets, which could form the basis for a repurposing screen. The graph in Figure 8 shows the drugs and other research compounds known to be active at the adenosine A1 receptor (*ADORA1*), one of the targets described above. The remaining drug repurposing candidates identified in the study are described further in the Appendix.

**Figure 8:**
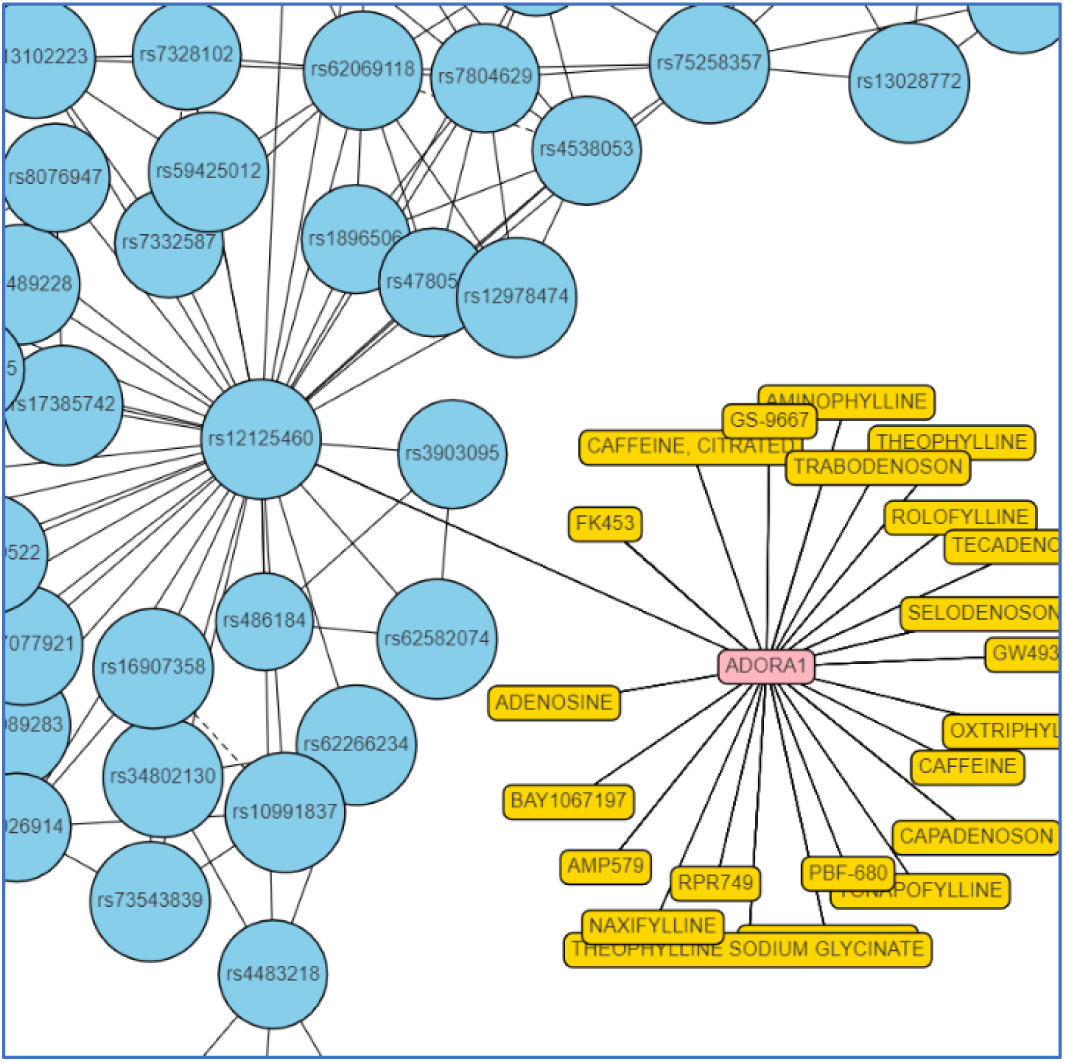
Network showing SNP signatures, risk-associated genes, and existing drug options in a sub-population of the sepsis disease architecture around a cluster containing an *ADORA1* related SNP.

### COVID-19 OVERLAP

For the signatures observed in sepsis and severe COVID-19 patients there are a number that are specific to sepsis, but the majority (43 of 71 - 61%) are shared both in sepsis and severe COVID-19 patients.

**Figure 9:**
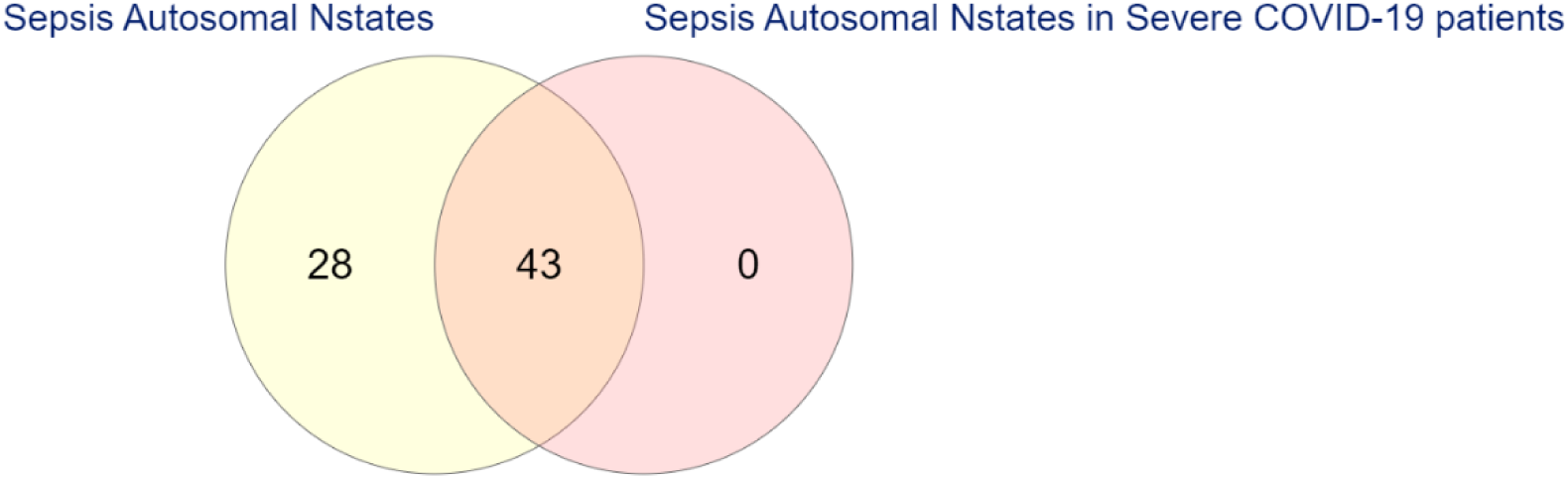
Venn diagram showing overlap of n-state signatures identified as sepsis risk factors in severe COVID-19 patients.

We found 33 sepsis disease signatures composed of autosomal SNPs that were present in patients who tested positive for COVID-19 and showed severe symptoms but were not observed in any COVID-19 positive patients showing mild symptoms or any sepsis controls.

One of the SNP genotypes combinations (signatures) found in both sepsis and severe COVID-19 patients, but not in sepsis controls or patients with mild (non-hospitalized) COVID-19 disease, contained three SNPs that mapped to six genes (Table 2). Two of the SNPs have been previously linked to cardiovascular and hypertension in phenotype-wide association studies (PheWAS) studies^47,48^ and functional annotations of the genes revealed that several of them are involved in neutrophil degranulation, leukocyte activation and immune effector processes. This disease signature was also observed in only 1,137 of the other 480,000 UK Biobank participants who are likely to suffer from complications in their immune responses because of this.

**Table 2:** Table showing SNP genotypes in a sepsis disease signature found both in sepsis and severe COVID-19 patients, but not in mild COVID-19 patients, with risk-associated SNPs and mapped genes.

| Autosomal SNP Genotypes | Sepsis + Severe COVID-19 SNPs | Gene Mappings |
| --- | --- | --- |
| Homozygous Minor Allele (2) | rs12026838 | SMYD3 |
| Homozygous Major Allele (0) | rs11924446 | CNST |
| Heterozygous (1) | rs117984853 | ACPP |
|  |  | DNAJC13 |
|  |  | AL357992.1 |
|  |  | AL031056.1 |

We will continue investigating the likely functional impacts of all the sepsis disease signatures found in severe COVID-19 patients in a further study. As more clinical and COVID-19 data become available in UK Biobank and other patient data sources, we will be able to analyze the clinical impact of these disease signatures in patients.

**Figure 10:**
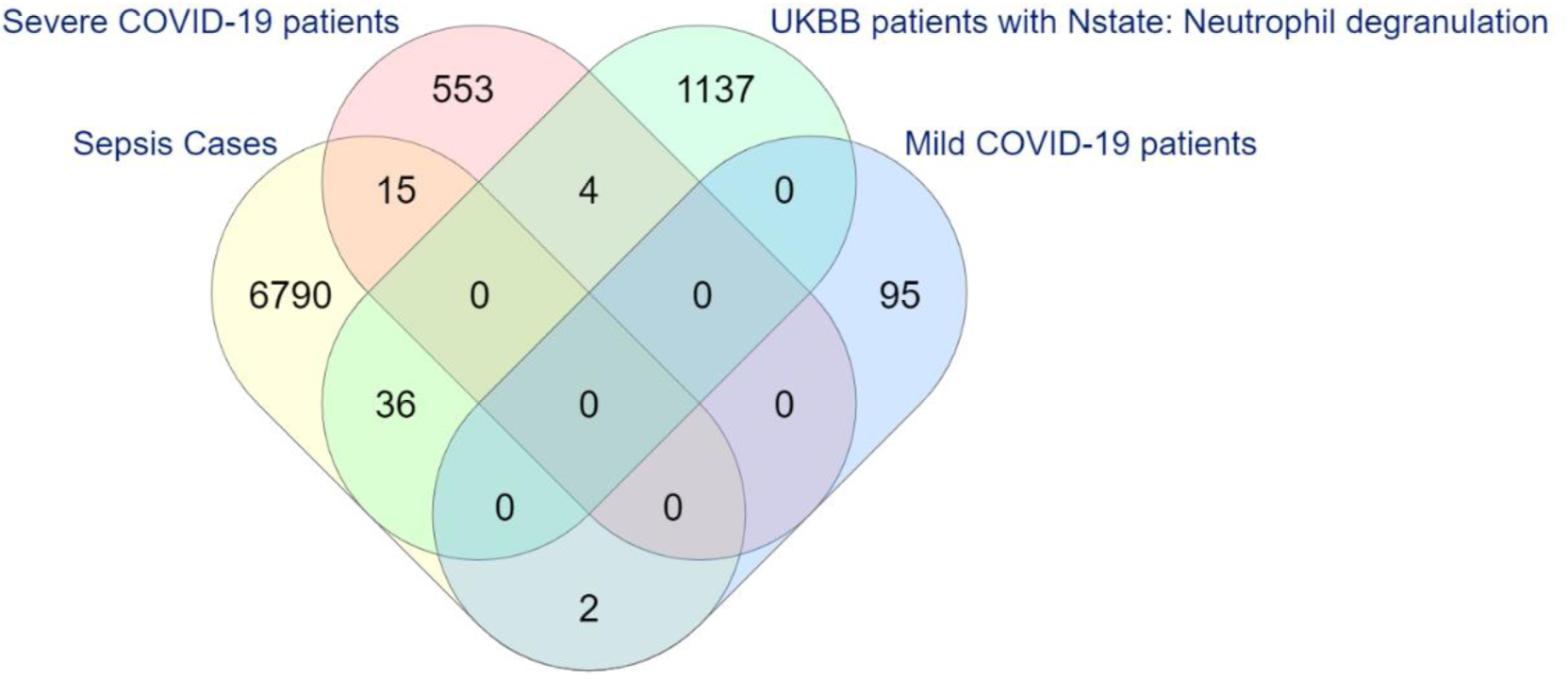
Venn diagram showing overlap of the UK Biobank participants that have the sepsis disease signature implicated in neutrophil degranulation, leukocyte activation and immune effector processes in the sepsis cases, severe COVID-19 patients and mild COVID-19 patients.

## Conclusion

Using only genetic data from the UK Biobank, we have identified 70 sepsis risk-associated genes that would not have been found using conventional GWAS approaches using the same dataset. We have further identified 59 drug repurposing candidates for 13 of these targets that have the potential to be directly effective in improving survival rates in sepsis. We found several other genes with strong mechanism of action hypotheses connected to existing drug targets that may form the basis of more novel drug discovery strategies.

We will be using these new insights into the disease to investigate several novel therapeutic strategies that may help to reduce the high mortality rates currently observed in patients who develop sepsis both within and without the context of COVID-19.

We continue to investigate the overlap between these sepsis-associated genetic signatures and those seen in COVID19-positive patients who present with life-threatening symptoms, including the development of viral sepsis. As more data from COVID-19 patients becomes available this may help to gain greater understanding into the mechanisms of late-stage disease and apparent clinical differences in patient responses to the SARS-CoV-2 virus.

New datasets may help elucidate links to observed epidemiological risk factors such as ethnicity, blood group and smoking. As these data become available for direct study, they may also provide new directions to search for effective therapies for patients outside of the antiviral space based on a better understanding of the host responses.

## Data Availability

All data will be made available in supplemental files in future versions of this paper and can be obtained by emailing

## Appendix

### Dataset Generation Details

#### CASE CRITERIA

Cases to <u>include</u> at least one of the following ICD-10 codes:

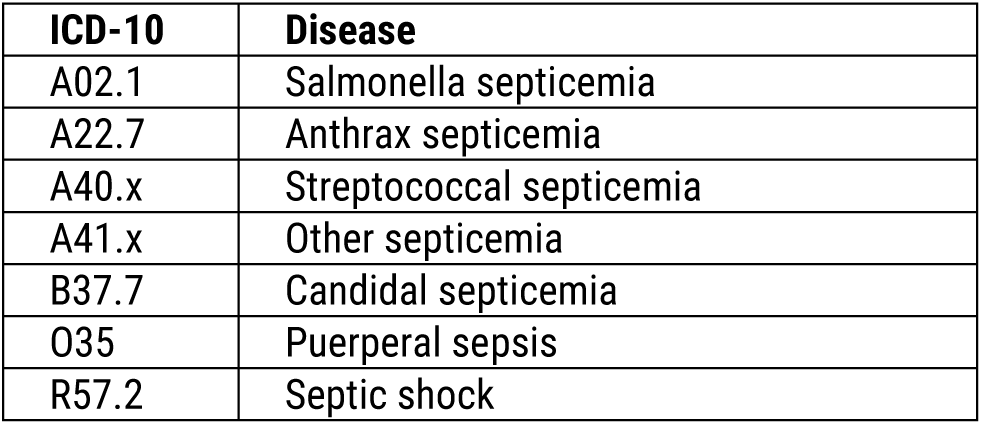

#### CONTROL CRITERIA

1. Controls to <u>exclude</u> any patients with the following ICD-10 codes:

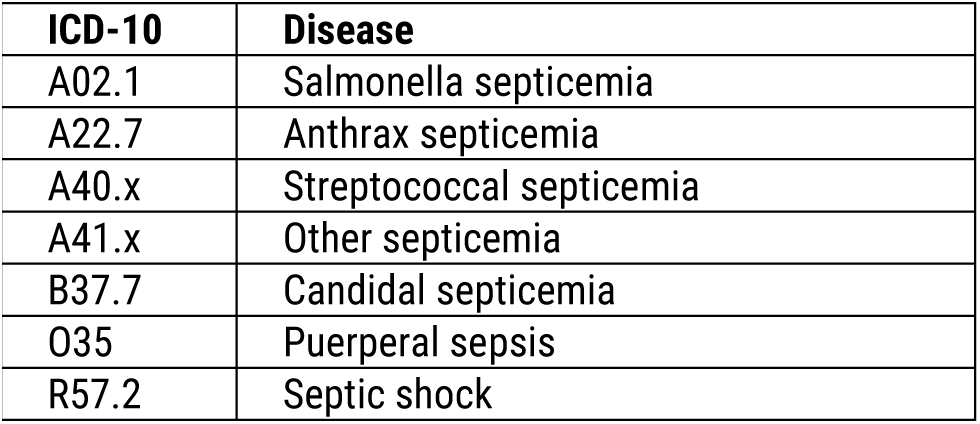
2. Controls to <u>include</u> at least one of the following ICD-10 codes:

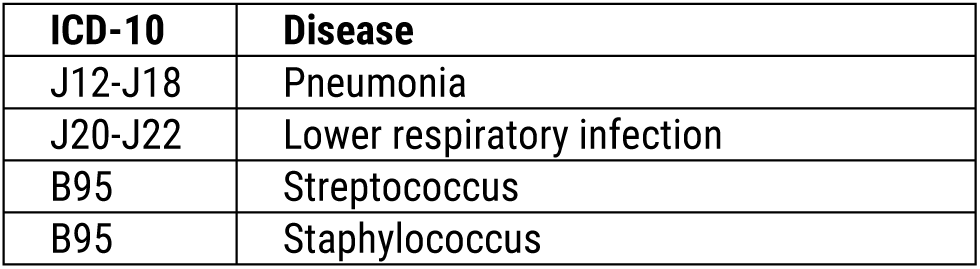
3. Controls to <u>include</u> least one of the following ICD-10 codes:

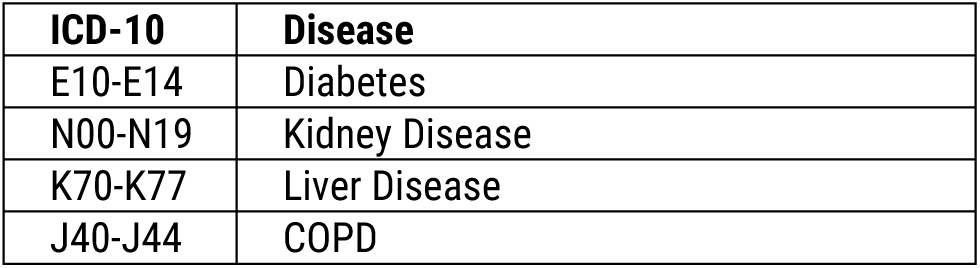

**Table 3.**
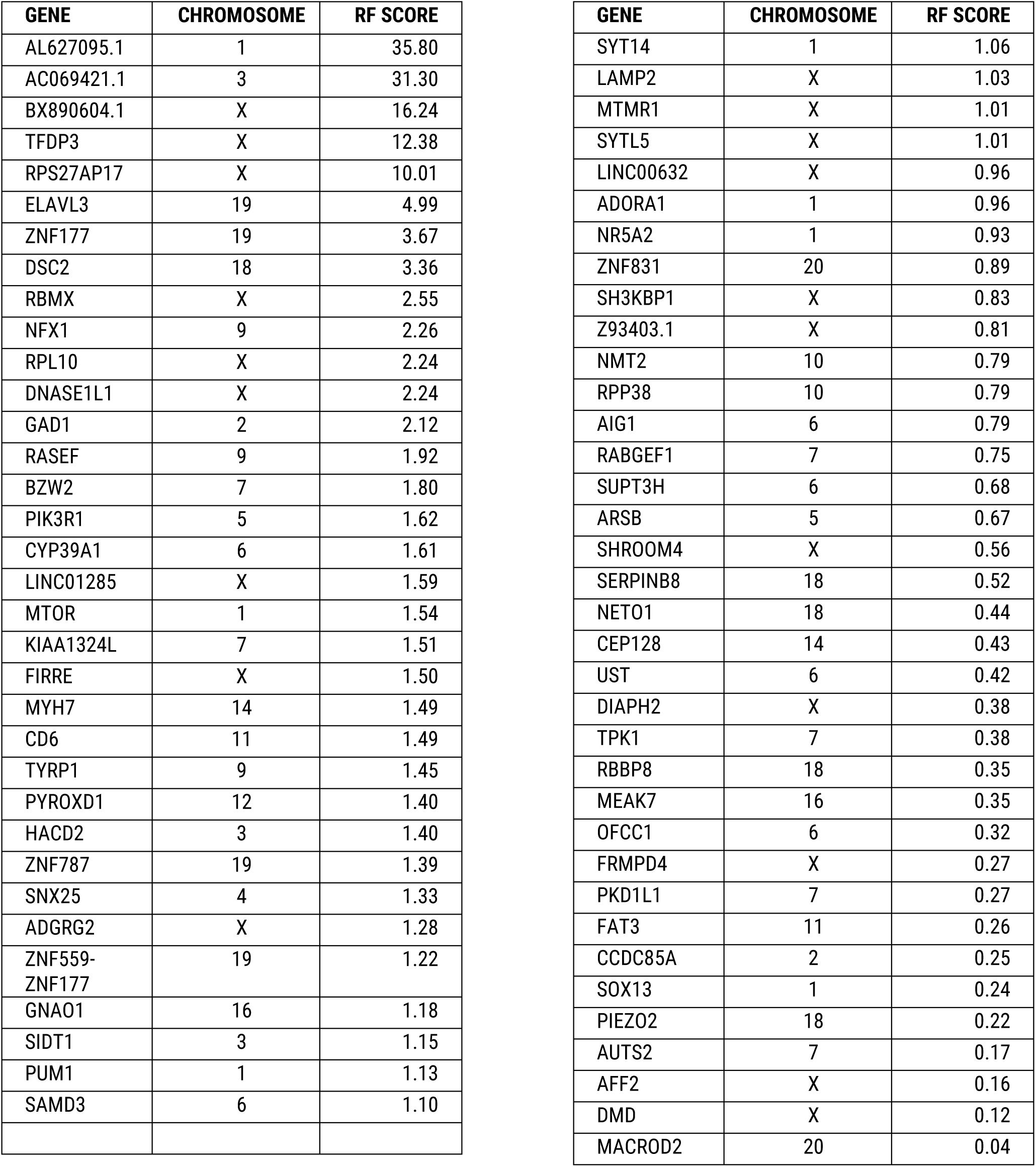
Table showing sepsis associated genes with chromosomal location and RF score

**Table 4.**
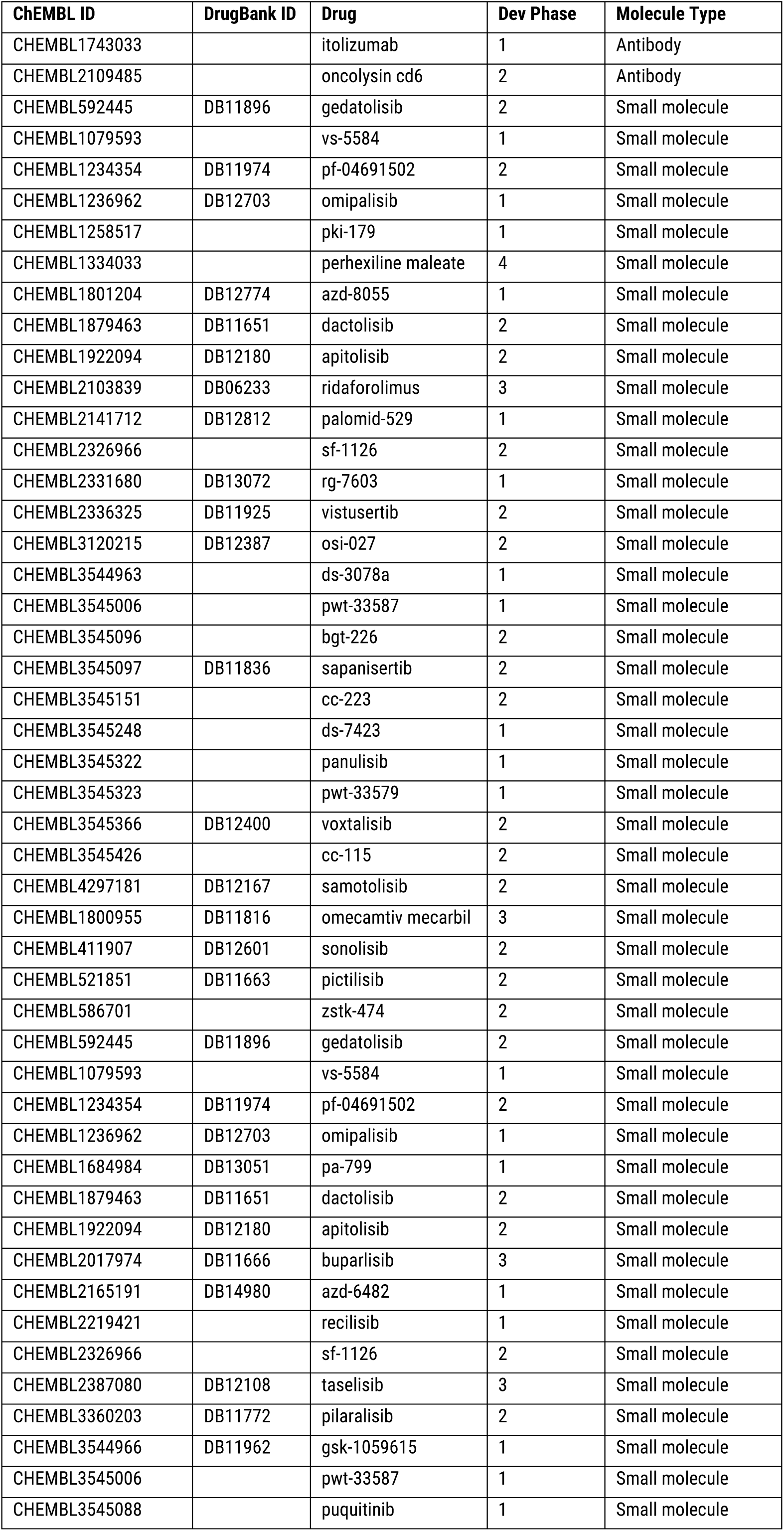
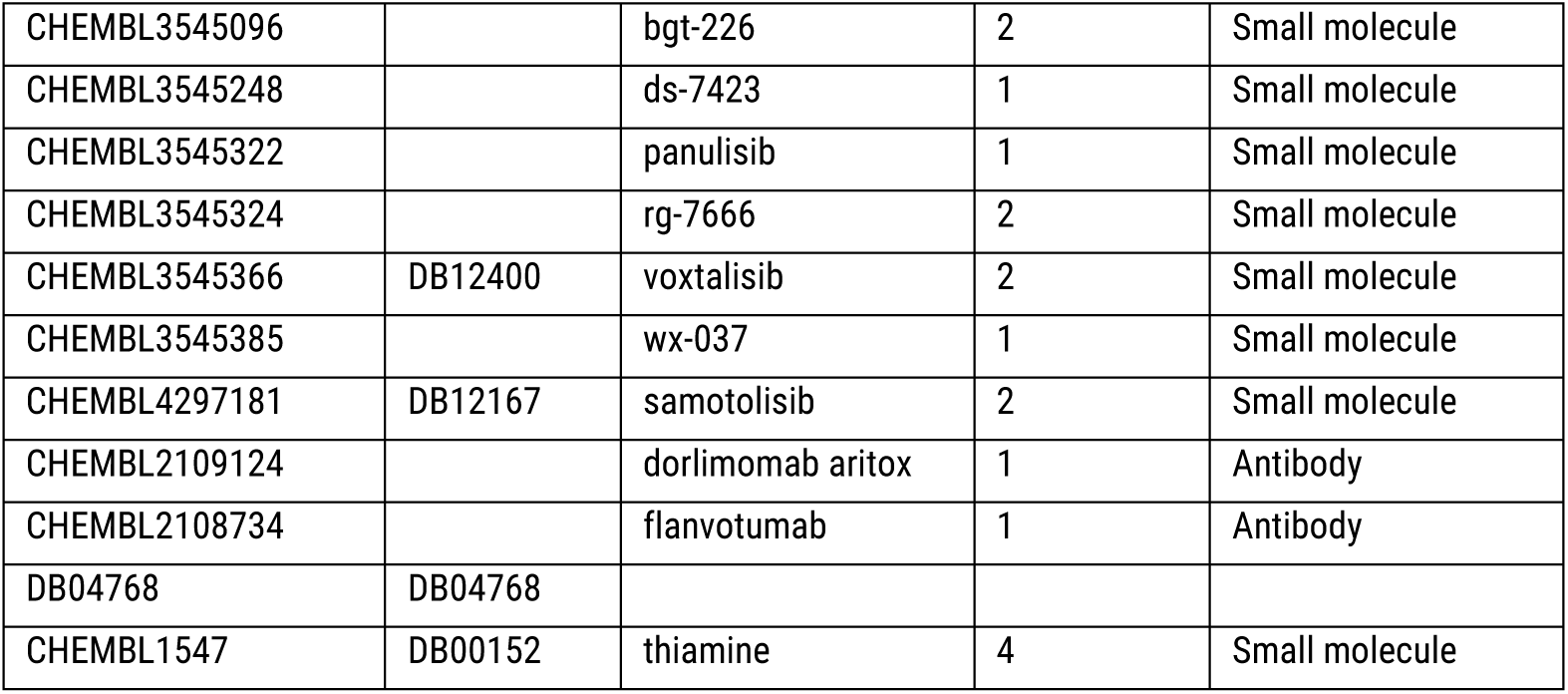
Table showing drug repurposing candidates for 13 target genes identified as being sepsis and COVID-19 related

#### Mining Terminology and Example

The overall process of mining, validation and scoring is shown below.

**Figure 11:**
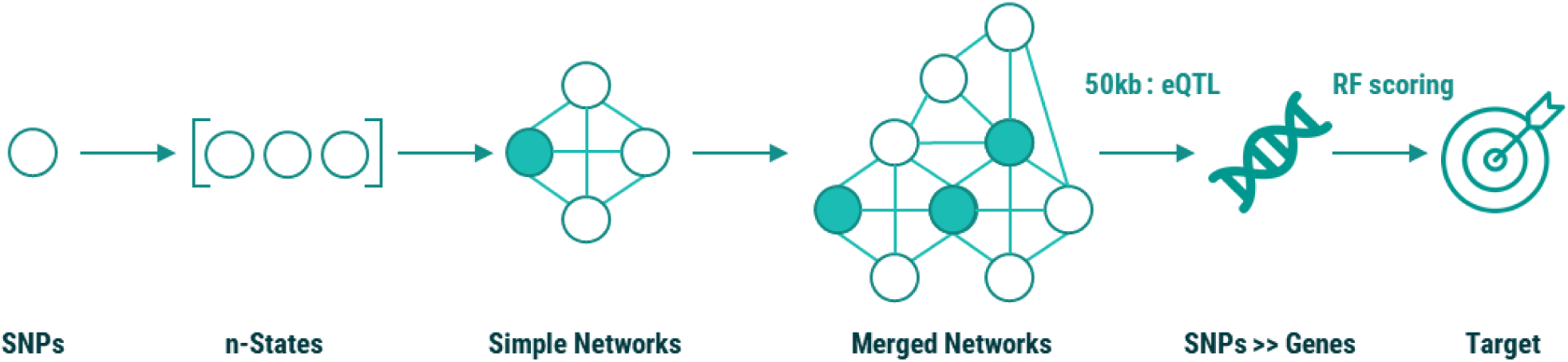
Stages of the **precisio**nlife mining, scoring and analysis process

In the small dataset approach 1,000 cycles of fully random permutation are used in the validation of the simple networks.

### GLOSSARY

n-states: :combinations of SNP genotypes (or other markers) significantly overrepresented in cases vs controls
Simple networks: :a set of n-states sharing at least one SNP genotype (the “critical SNP genotype”). These networks are validated with a certain FDR threshold, and sometimes known as “validated simple networks”.
Critical SNP genotypes: Merged networks: :validated SNP genotypes (SNP genotypes defining validated simple networks) the union of n-states from one or more networks

